# Wearable-based digital biomarker provides a valid alternative to traditional clinical measures for post-stroke upper-limb motor recovery

**DOI:** 10.1101/2025.01.13.25320461

**Authors:** Ryan Wang, Catherine E. Lang, Mary E. Stoykov, Paolo Bonato, Sunghoon I. Lee

## Abstract

Existing clinical assessments for upper-limb motor rehabilitation post-stroke pose limitations as endpoints for clinical trials. This study aims to develop a wearable-based digital biomarker for assessing motor recovery using accelerometer data collected in naturalistic environments. The study analyzed approximately 23,000 hours of data from 215 participants, including subacute and chronic stroke survivors and healthy individuals. A novel analytical approach decomposed continuous accelerometer data into a lower-level unit of motor behaviors called movement segments, from which key features were extracted and aggregated using a linear mixed-effects model to produce a composite biomarker. The resulting digital biomarker demonstrated excellent interpretability, reliability, concurrent validity, discriminant validity, known-group validity, and responsiveness, enabling a nearly 66% reduction in the required sample size for clinical trials compared to traditional measures. These findings highlight its potential as a low-burden, scalable assessment tool for upper-limb motor recovery, with applications in both clinical trials and routine clinical practice.

## Introduction

The availability of valid, reliable, and responsive outcome measures is imperative to support the efficient and effective development of novel therapeutic interventions for upper-limb motor rehabilitation post-stroke [1, 2]. Currently, clinician-performed assessment tools, such as the Fugl-Meyer Assessment of the Upper Extremity (FMA-UE) and Action Research Arm Test (ARAT), which evaluate patients’ behavioral phenotypes within laboratory environments, are considered the gold standard for domain-specific endpoints [2]. However, they rely on ordinal scales with limited granularity and sensitivity [3], are affected by significant ceiling and floor effects [4], and are vulnerable to variability caused by external factors, such as patient motivation or assessors’ encouragement during motor tasks [5]. Furthermore, they demand substantial resources, including trained specialists for administration [6] and ongoing training to maintain reliability [7], which offers only episodic snapshots of a patient’s response trajectory. Critically, these tools fail to capture biologically meaningful phenotypes in ecologically valid settings, despite the time- and environment-dependent nature of neural recovery [8]. These shortcomings have also posed significant barriers to translating research findings into clinical practice, despite numerous completed trials [1, 2].

Digital biomarkers—defined as indicators of biological processes collected from digital technologies, including wearable devices [9]—have emerged as a promising alternative [10, 11]. In the realm of post-stroke upper-limb research, wearable-based outcome measures built on continuous inertial data during patients’ performance of everyday activities could support low-cost, objective, and longitudinal assessment of the motor recovery process [3, 6]. However, since their inception in the early 2000s [12], these approaches have predominantly focused on a unidimensional aspect of motor performance—*limb activity levels*—which quantify wrist acceleration magnitudes of the stroke-affected limb or compare them to the contralateral limb [13]. While simple, easy to interpret, and exhibiting solid convergent validity, these methods have demonstrated weaker concurrent validity and responsiveness compared to standardized domain-specific endpoints, likely due to their inability to capture the multidimensional complexity of upper-limb motor behaviors. Consequently, clinician-performed assessments remain the primary choice for endpoints in clinical trials [2].

To bridge these gaps, we investigate a new composite digital biomarker for assessing stroke upper-limb motor recovery, constructed using accelerometer data from the stroke-affected wrist and a set of machine-learning algorithms. However, extracting information about biologically meaningful phenotypes from unstructured and naturalistic upper-limb movements poses significant challenges due to the wide variation in the types of ADLs and the diverse ways in which these ADLs are executed, both across and within individuals [13]. To that end, we employed a novel data analytic method, motivated by theories of motor control and behavior [14–17], which decomposes patients’ performed linear movements into a lower-level unit of motor behavior called *movement segments* [18, 19]. By focusing its analysis on these isolated segments of linear movements, the proposed technique could capture meaningful phenotypes, irrespective of the amount or type of ADLs undertaken. Our central hypothesis is that kinematic and temporal characteristics of these movement segments are indicative of upper-limb motor recovery post-stroke and can be used to construct a valid, reliable, and responsive digital biomarker.

The proposed digital biomarker, according to the BEST biomarker category [20], is a monitoring biomarker that can be repeatedly used to assess upper-limb motor recovery in stroke survivors. Its primary application is to serve as a cost-effective endpoint in clinical trials. However, it also holds potential for use in clinical care to bolster more personalized therapy [2, 3] and patients’ self-managed rehabilitation [21].

## Results

### Overview

We employed a data-driven approach (Fig. 1A) to construct the proposed digital biomarker, leveraging continuous inertial data collected by a three-axis accelerometer placed on the stroke-affected wrist. The biomarker was developed and validated using approximately 23,000 hours of accelerometer data from 215 individuals in their naturalistic environments [22]. The dataset [22–24] included three different subpopulations captured across three independent studies, as summarized in Fig. 1B: 61 subacute-stage stroke survivors [25], 79 chronic-stage stroke survivors [26, 27], and 75 neurologically-intact, community-dwelling adults [28, 29].

**Fig 1.**
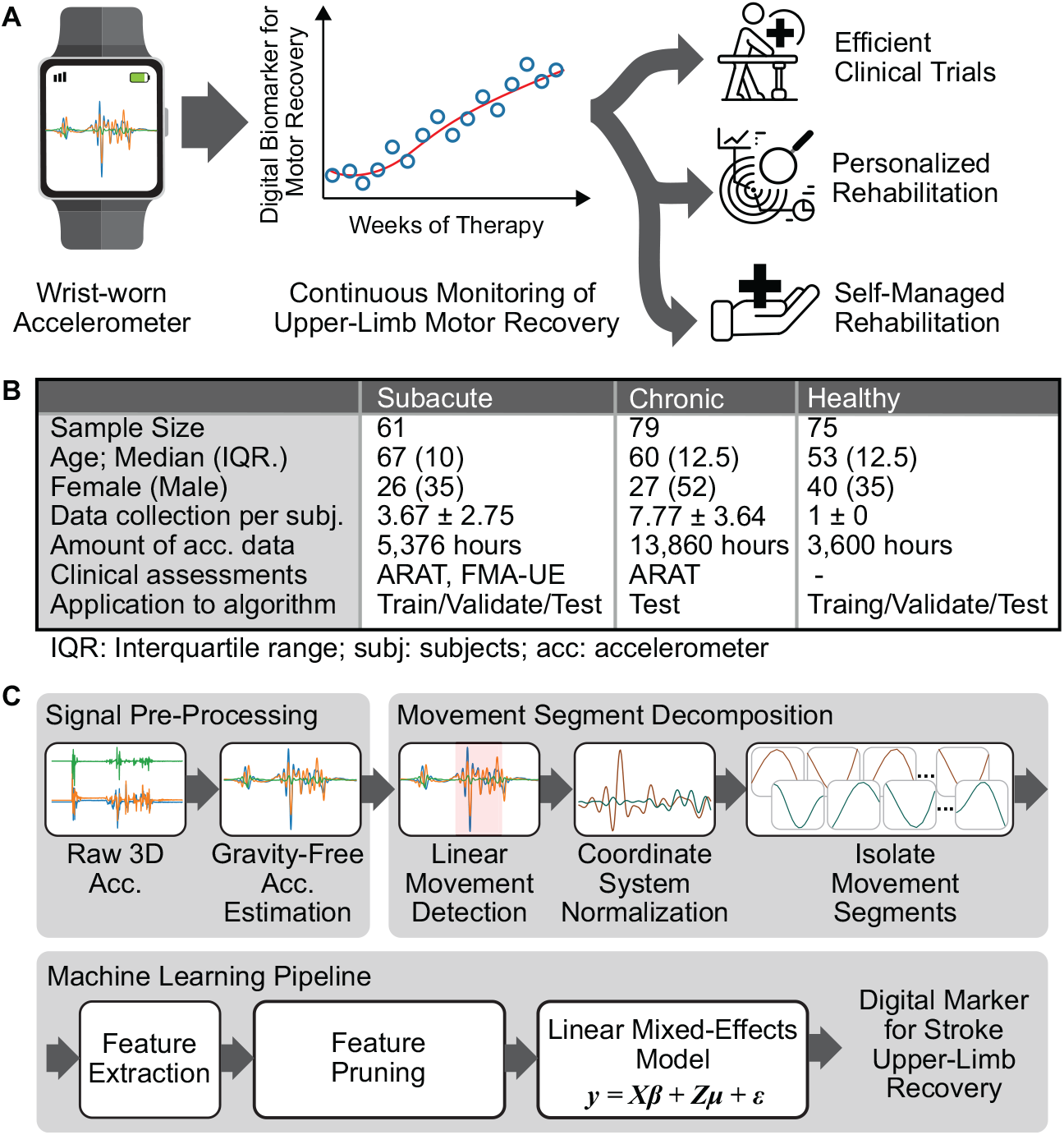
Overview of the proposed digital biomarker and study design. **A** Graphical representation of our system, where wrist-worn accelerometer data were utilized to construct a digital biomarker for post-stroke motor recovery. **B** Summary of the three datasets used for the biomarker’s development and validation. **C** Flowchart of the biomarker development process: accelerometer data were first preprocessed using a band-pass filter to attenuate gravitational components and sensor noise. Linear movements were then identified, and movement segments were decomposed. The machine learning pipeline included feature extraction from movement segments, feature pruning, and training of a linear mixed-effects model.

Subacute and chronic stroke survivors participated in multiple assessments. Subacute participants, receiving conventional rehabilitation therapy, were assessed up to eight times over 24 weeks, while chronic participants, enrolled in an eight-week upper-limb rehabilitation clinical trial, completed up to 14 assessments over 20 weeks. Each assessment began with clinical evalautions: subacute participants completed ARAT and FMA-UE, while chronic participants completed only ARAT. Continuous accelerometer data from the stroke-affected limb during the following 24 hours of naturalistic activities were collected and analyzed. Healthy individuals underwent a single cross-sectional session, with accelerometer data from both wrists included in the analysis. They were assigned with the highest possible clinical scores without administrating the assessments, indicating full motor capacity and no impairment.

### Development and validation of the digital biomarker

Fig. 1C visually summarizes the data analytic pipeline, which consists of acceleration data preprocessing, movement segment decomposition, and machine learning model training, as previously described [18]. Briefly, a band-pass filter with cutoff frequencies of 0.25–2.5 Hz was applied to attenuate gravitational effects, motion artifacts, and sensor noise [25, 27, 28]. A heuristic algorithm then identified *linear movements*, indicating a relatively straight path within 3D space [30]. Linear movements were decomposed into movement segments based on velocity zero-crossings within the two primary axes of motion and clustered into three groups as described in prior work [19]: 1) the homogeneous set, with smooth, bell-shaped velocity profiles typical of neurologically intact individuals [31]; 2) the outlier set, featuring irregular morphologies indicative of impairment-specific motor characteristics [17]; and 3) the entire set, encompassing both for comprehensive motor characterization. Kinematic, morphological, and temporal features relevant to stroke-related motor deficits were extracted from each cluster and aggregated at the subject level using statistical functions (see Methods for details) [19].

The machine learning algorithm was designed to integrate relevant features into a comprehensive indicator of motor status, ensuring that the resulting composite biomarker satisfies core statistical validation criteria: reliability, concurrent validity, and responsiveness [32, 33]. First, feature pruning was performed to eliminate unreliable and irrelevant features. A linear mixed-effects regression model was then applied where 1) fixed effects captured cross-sectional associations with min-max-normalized ARAT scores, supporting concurrent validity, and 2) random effects modeled within-subject temporal correlations, addressing responsiveness. The resulting biomarker primarily ranged from 0 to 1, with lower values indicating more severe motor impairment, though its theoretical range was unbounded and thus free from floor or ceiling effects.

The model was trained using a combined dataset of subacute stroke survivors and healthy subjects, as subacute participants exhibited more dynamic changes in motor status over time than chronic participants, which was essential for developing a responsive biomarker. Healthy subjects were included to represent neurologically intact phenotypes. We utilized nested leave-one-subject-out cross-validation (LOSOCV) for evaluation so that no identical subject data was included in the training, hyperparameter tuning, and testing processes [32]. We used chronic data to demonstrate the biomarker’s generalizability beyond the subacute stage. In this case, the model trained and validated on subactue and healthy data through LOSOCV was tested on the chronic data.

### Validation of the digital biomarker in subacute stroke survivors

The proposed biomarker for upper-limb motor severity demonstrated excellent test-retest reliability in subacute stroke survivors [34], with an ICC(3,1) of 0.93, as shown in Fig. 2A. Reliability was assessed by splitting each individual’s 24-hour accelerometer data into two groups, alternating between even- and odd-numbered 30-minute time windows, and independently computing the biomarker from each group. We also estimated two key metrics for measurement errors—Standard Error of Measurement (SEM) and Minimum Detectable Change (MDC) [35]—and compared them with clinician-performed endpoints in Table 1. The biomarker’s SEM and MDC marked a 38-52% reduction compared to the min-max-normalized ARAT and FMA-UE, indicating greater sensitivity to change.

**Table 1.** Comparison of MDC and SEM for the Proposed Biomarker, ARAT, and FMA-UE.

| Metric | Measurement | Subacute | Chronic |
| --- | --- | --- | --- |
| SEM | Proposed Biomarker | 0.036 | 0.021 |
|  | Normalized ARAT | 0.074 | 0.042 |
|  | Normalized FMA-UE | 0.059 | – |
|  | Actual ARAT | 4.25 | 2.42 |
|  | Actual FMA-UE | 3.91 | – |
| MDC | Proposed Biomarker | 0.10 | 0.058 |
|  | Normalized ARAT | 0.21 | 0.12 |
|  | Normalized FMA-UE | 0.16 | – |
|  | Actual ARAT | 11.77 | 6.70 |
|  | Actual FMA-UE | 10.83 | – |

**Fig 2.**
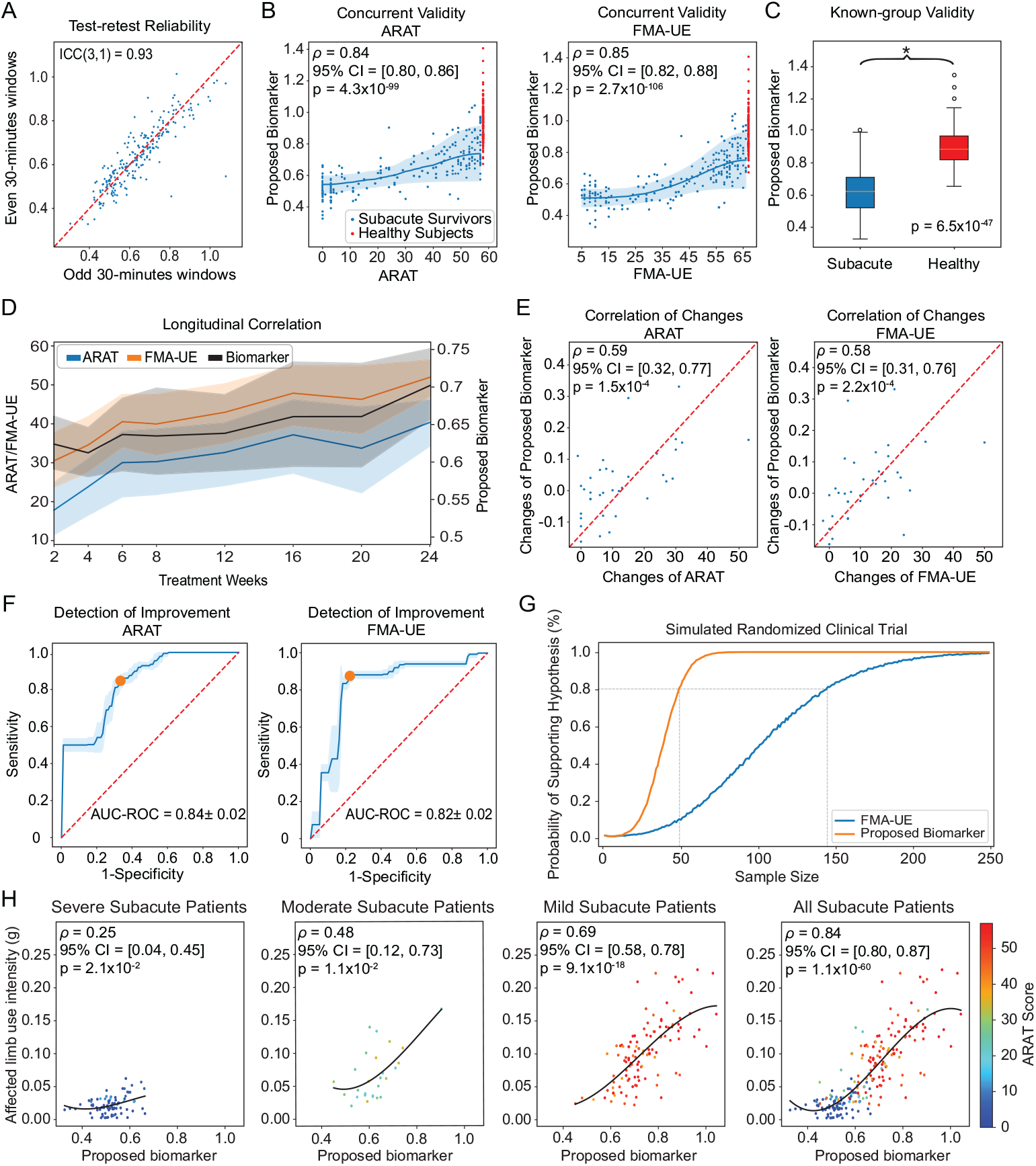
Validation of the digital biomarker in subacute stroke survivors. **A** Scatter plot assessing test-retest reliability between even-numbered (y-axis) and odd-numbered (x-axis) time windows across all clinical visits. **B** Scatter plots showing the correlation between the proposed biomarker (y-axis) and standardized clinical assessments (x-axis), including the mean trend (solid line) and two standard deviations (shaded area). **C** Boxplot comparing the distribution of the biomarker between subacute-stage stroke survivors and healthy controls, with statistical significance assessed using the Wilcoxon rank-sum test. **D** Temporal trends of the proposed biomarker and clinical measures over the treatment weeks, showing the mean trend and two standard deviations. **E** Scatter plots illustrating correlations between changes in the proposed biomarker (y-axis) and changes in clinical assessments (x-axis). **F** Receiver operating characteristic curves demonstrating the proposed biomarker’s ability to detect meaningful improvement in clinical assessments, with mean and one standard deviation obtained via leave-two-subject-out bootstrapping. The orange dots represent the operation points that maximize F1 scores for further analysis. **G** Graphical representation of a simulated clinical trial, showing the relationship between the probability of supporting the hypothesis (y-axis) and required sample size (x-axis). **H** Scatter plots depicting the correlation between the proposed biomarker (x-axis) and affected limb use intensity (y-axis) across severity groups for subacute stroke survivors. Colors indicate functional capacity measured by ARAT, with warmer colors representing greater capacity.

Fig. 2B demonstrates the biomarker’s strong concurrent validity against standardized endpoints. The Spearman’s rank correlation coefficients (ρ) were 0.84 with ARAT (*p* = 4.3 × 10^−99^, 95% confidence interval (CI) = [0.80, 0.86]) and 0.85 with FMA-UE (*p* = 2.7 × 10^−106^, 95% CI = [0.82, 0.88]), indicating excellent agreements [36]. Additionally, the biomarker avoids ceiling or floor effects due to its theoretically infinite score range, unlike ARAT and FMA-UE [4].

Fig. 2C demonstrates the biomarker’s strong known-group validity, effectively distinguishing healthy behavioral phenotypes from those of subacute participants (*p* = 6.5 × 10^−47^; Wilcoxon rank-sum test), as hypothesized. Extended Data Fig. 1 provides a detailed illustration of the digital biomarker distributions for the two populations, showing a value range of 0.33 to 1.04 in subacute patients and 0.67 to 1.41 for healthy individuals. We also assessed the biomarker’s discriminant validity against demographic factors—specifically age, gender, and whether the dominant or non-dominant limb was affected—based on the hypotheses that the biomarker should remain unaffected by the associated motor characteristics. The biomarker showed weak correlations with age in both healthy and subacute participants, as shown in Extended Data Fig. 2. Additionally, no significant differences were observed between genders in either healthy or subacute groups (Extended Data Fig. 3A, B), nor between affected dominant and non-dominant limbs among subacute participants (Extended Data Fig. 4A).

The responsiveness of the biomarker was examined using three different approaches: 1) correlation between the temporal trajectories of the proposed biomarker and standardized assessments, 2) correlations between changes in biomarker scores and changes in standardized measures, and 3) dichotomous detection of improvement defined based on standardized measures. Fig. 2D shows that the biomarker exhibits a strong agreement to standardized measures for population-level trajectory over the 24-week study period. Data were grouped by collection time (i.e., weeks from baseline), and the mean trajectories with two standard deviations were visualized for all measures. The mean trajectory of the biomarker exhibited a Spearman correlation of 0.93 with ARAT (*p* = 8.6 × 10^−4^, 95% CI = [0.65, 0.99]) and 0.95 with FMA-UE (*p* = 2.6 × 10^−4^, 95% CI = [0.75, 0.99]). Fig. 2E shows that changes in the proposed biomarker between the first and last visits exhibited Spearman correlation coefficients of 0.59 with ARAT changes (95% CI = [0.32, 0.77], ρ = 1.5 × 10^−4^) and 0.58 with FMA-UE changes (95% CI = [0.31, 0.76], ρ = 2.2 × 10^−4^), demonstrating excellent longitudinal agreement [37].

Fig. 2F illustrates the biomarker’s capability to detect motor improvements based on external criteria. Participants were classified as responders if the difference in their clinical scores between the first and last visits exceeded the MDC specified in Table 1, and as non-responders otherwise. Of the 61 participants, 36 had more than one assessment session, with 14 were identified as responders based on ARAT’s MDC and 17 based on FMA-UE’s MDC. The biomarker achieved an Area Under the Receiver Operating Characteristic Curve (AUC-ROC) of 0.84 ± 0.02 for ARAT and 0.82 ± 0.02 for FMA-UE, both indicative of strong detection performance [37]. We further analyzed the misclassified samples at an operating point on the ROC that maximized the F1 scores (indicated by dots in Fig. 2F). We observed that, among the false positive samples, five out of six in ARAT and all three in FMA-UE were influenced by floor and ceiling effects (i.e., scores near the minimum on the first visit or near the maximum on the last visit). For false negatives, two of three ARAT cases had score improvements comparable to the MDC, suggesting potential false improvements. These findings highlight the biomarker’s potential to mitigate the limitations of traditional clinical measures in accurately determining patient responses.

### Hypothetical clinical trial

Based on the statistical properties of our digital biomarker, we evaluated its potential advantages over a traditional clinician-performed endpoint (i.e., FMA-UE) in a simulated randomized clinical trial. We employed the statistical framework based on Monte Carlo simulation, proposed by Mori *et al*. [11], for structuring this hypothetical trial. It was designed to examine the long-term effect of constraint-induced movement therapy versus conventional therapy among subacute stroke survivors, drawing upon findings from a randomized clinical trial conducted by Boake *et al*. [38]. Fig. 2G shows that the choice of endpoints significantly influences the required sample size to achieve the desired level of statistical power. For instance, achieving an 80% probability of supporting the hypothesis requires 143 participants per arm with the FMA-UE, compared to only 48 with the proposed digital biomarker, marking a 66.0% reduction. We attribute this reduction to the substantially smaller SEM of the proposed biomarker, as shown in Table 1. These findings suggest that the proposed digital biomarker could serve as a valid and efficient endpoint, potentially reducing the cost and sample size of clinical trials.

### Comparison between the proposed biomarker and the affected limb use intensity

The biomarker was compared to the activity intensity of the stroke-affected limb, computed by accumulating the acceleration magnitude throughout the monitoring period [13, 39]. While affected limb intensity showed a strong correlation with the digital biomarker (ρ = 0.84, 95% CI = [0.80, 0.87]), as well as with ARAT (ρ = 0.82, 95% CI = [0.79, 0.85]) and FMA-UE (ρ = 0.84, 95% CI = [0.81, 0.87]), its correlation with the biomarker varied by stroke severity. Specifically, the correlation was minimal in participants with severe motor severity and became more pronounced in participants with moderate to mild motor severity, as illustrated in Fig. 2H. Severity groups were defined using ARAT scores [40]. This relationship aligns with the threshold hypothesis proposed by Lucena *et al*. [41], which suggests that stroke survivors generally refrain from using their impaired limb until reaching a certain level of recovery [42]. This adds to the findings supporting that the proposed biomarker captures informative recovery phenotypes—e.g., more closely tied to functional ability (ARAT) and motor impairment (FMA-UE)—rather than merely limb activity.

### Interpretation of the digital biomarker via feature analysis

Table 2 summarizes the key features that constitute the proposed biomarker to interpret its content validity. Descriptive functions represent statistical properties derived from movement segments, while aggregation functions describe how these descriptive features were synthesized at the subject level. Because different features were used to train the machine learning model in each outer LOSOCV iteration, we identified a set of features that were selected 100% of the folds and then selected representative features among features with high inter-feature correlations [19].

**Table 2.**
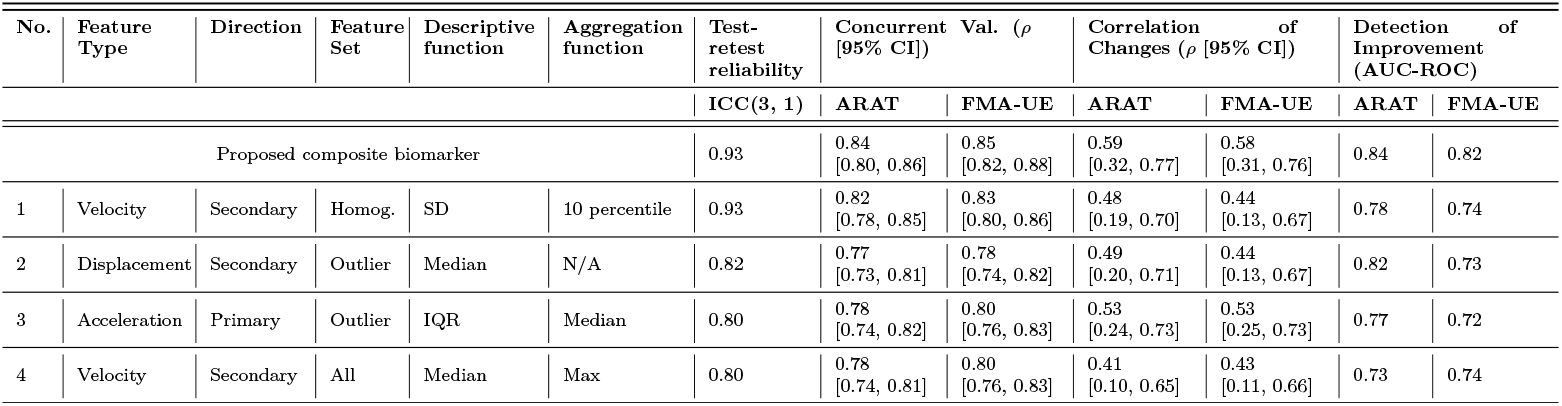
Key Features Contributing to the Digital Biomarker. Summary of the descriptions and clinimetric properties of the key features. ‘Feature Type’ denotes the source data from which the features were extracted. ‘Direction’ specifies the axis of linear movement associated with each feature. ‘Descriptive functions’ capture statistical properties derived from the time series of individual movement segments, while the ‘aggregation functions’ summarize these descriptive features at the subject level. For features utilizing only descriptive or aggregation functions, the inapplicable function was marked as N/A (Not Available).

These features captured distinct aspects of motor performance, utilizing acceleration, velocity, and displacement data across homogeneous, outlier, and entire movement segments in both primary and secondary directions. Features 1 and 3 reflect kinematic variance in linear movements, indicating that as motor severity worsens, participants generate more restricted kinematics, resulting in reduced variance [18, 19, 42]. Feature 2 captures the typical distance of linear movements, showing that displacement decreases with increasing motor severity [15–17]. Lastly, Feature 4 represents maximal motor performance (i.e., velocity), which contributes to the digital biomarker’s strong association with clinician-rated measures of maximal functional ability (ARAT) and impairment (FMA-UE) [43]. Though each feature independently showed acceptable reliability, concurrent validity, and responsiveness, the composite biomarker—integrating these multidimensional features—outperformed in all criteria.

### Validation of the digital biomarker in chronic stroke survivors

The digital biomarkers for chronic stroke survivors were generated by applying the machine learning model, trained and validated on the combined subacute and healthy datasets, to the chronic dataset. The proposed biomarker showed excellent reliability in the chronic data, with an ICC(3,1) of 0.96 (Fig. 3A). The SEM and MDC of the digital biomarker were 0.021 and 0.058, respectively, both significantly smaller than those of the clinical assessment (Table 1).

**Fig 3.**
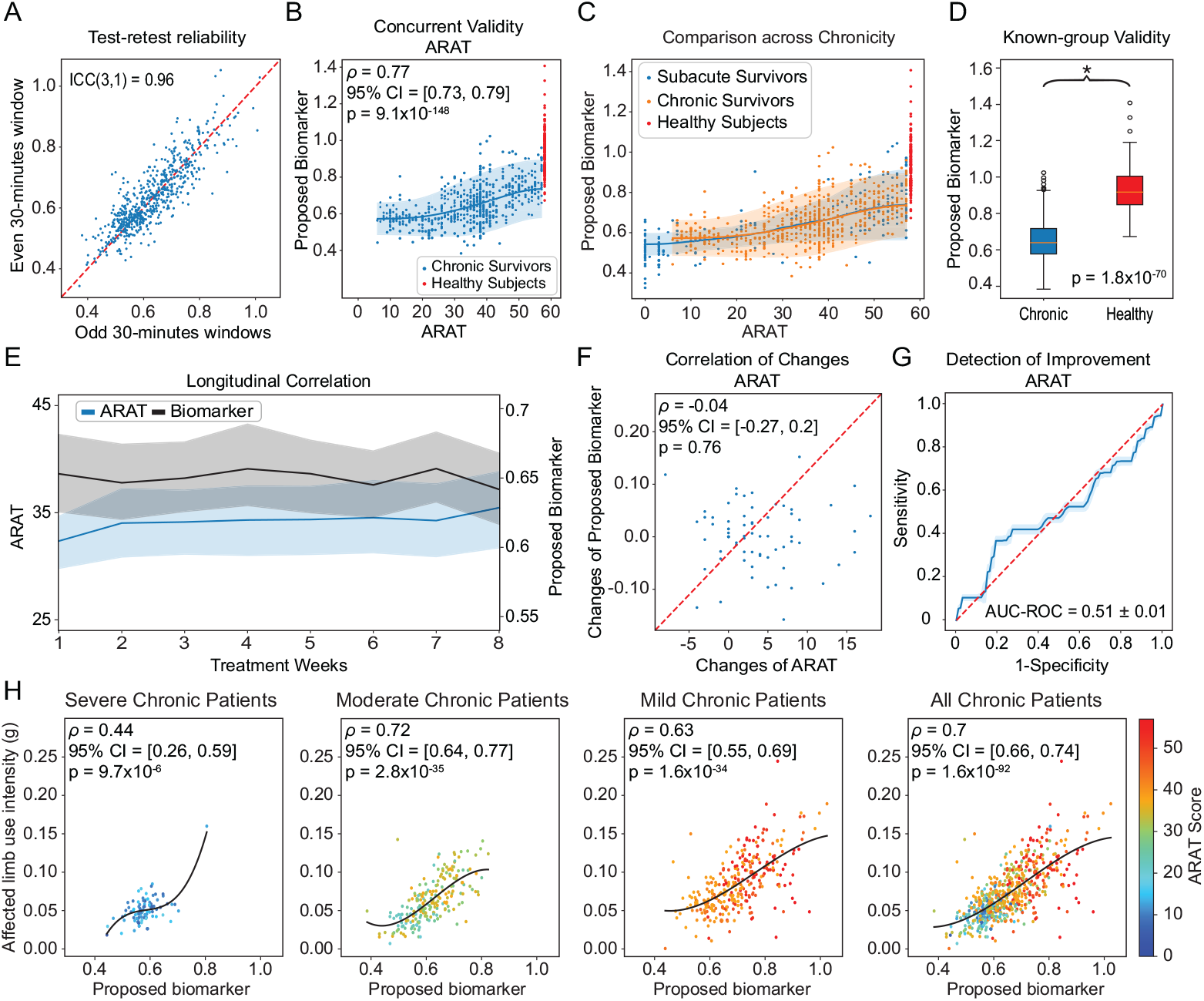
Validation of the digital biomarker in chronic stroke survivors. **A** Scatter plot investigating test-retest reliability between even-numbered (y-axis) and odd-numbered (x-axis) time windows across all clinical visits. **B** Scatter plot showing the proposed biomarker (y-axis) and the ARAT assessments (x-axis). the correlation between the proposed biomarker (y-axis) and ARAT (x-axis). **C** Scatter plot comparing the distribution of the proposed biomarker across subacute (blue), chronic stroke survivors (orange), and healthy subjects (red) against their ARAT assessments. **D** Boxplot comparing the distribution of the biomarker between chronic-stage stroke survivors and the healthy control group, with statistical significance assessed using the Wilcoxon rank-sum test. **E** Temporal trend of ARAT assessment and the proposed biomarker over the treatment weeks, including the mean trend and two standard deviations. **F** Scatter plot illustrating the correlation between changes in the proposed biomarker (y-axis) and changes in ARAT (x-axis). **G** ROC curve showing the proposed biomarker’s ability to detect meaningful improvement in clinical assessments, with mean and one standard deviation obtained via leave-two-subject-out bootstrapping. **H** Scatter plot depicting the relationship between the proposed biomarker (x-axis) and the affected limb use intensity (y-axis) across severity groups and for chronic stroke survivors. Colors indicate functional capacity measured by ARAT, with warmer colors representing greater capacity.

Fig. 3B supports the biomarker’s strong concurrent validity with ARAT [36], showing a Spearman’s correlation of 0.77 (*p* = 9.7 × 10^−148^, 95% CI = [0.73, 0.79]). The average trend line between the biomarker and ARAT for chronic participants aligned closely with that of the subacute population, as shown in Fig. 3C, suggesting the biomarker captures a similar construct across chronicity. However, the variance in the digital biomarker for a given ARAT score was larger for chronic participants, possibly influenced by individual-level factors, such as psychological, sociological, and environmental factors [25]. Fig. 3D demonstrates strong known-group validity between chronic participants and healthy individuals (*p* = 1.8 × 10^−70^; Wilcoxon rank-sum test). Extended Data Fig. 1 provides detailed value distributions for these groups. For discriminant validity, the biomarker showed weak correlations with age and no significant gender differences, as shown in Extended Data Figs. 2C and 3C, respectively. However, a significantly higher biomarker score was observed when the dominant limb was affected (Extended Data Fig. 4B). This difference potentially reflects chronic participants’ adaptation of the dominant limb in daily activities over time, leading to improved motor performance [44].

Fig. 3E, F, and G summarize the biomarker’s responsiveness in chronic participants. Unlike subacute participants, chronic participants showed minimal motor improvement from the intervention, as indicated by prior studies [26] and visualized in Fig. 3E. Furthermore, among 69 of the 79 chronic participants with multiple clinical visits, the average change in ARAT from baseline to the final treatment week was 4.7 ± 4.4, which was smaller than the MDC of 6.7 derived from our data. Similarly, the biomarker’s change was 0.044 ± 0.038, with an MDC of 0.058. These limitations significantly affected the ability to determine and detect meaningful changes in patient conditions, as shown in Fig. 3F and G. The relationship between the biomarker and affected limb intensity paralleled the findings in acute participants, reinforcing the threshold hypothesis, as shown in Fig. 3H.

### Influence of Compensatory Behaviors on Digital Biomarker

There is an ongoing debate among rehabilitation clinicians and researchers about whether wearable-based motor assessments should differentiate upper-limb movements performed with normative shoulder and elbow biomechanics from those involving compensatory or substitutive movements [13]. Although our digital biomarker was not explicitly trained to assess movement quality, results in Fig. 2D, E, and F indicate that it reliably detects changes (or lack thereof) in motor status, showing strong alignment with ARAT and FMA-UE scores, both of which penalize compensatory behaviors [45]. For example, Extended Data Fig. 5A compares chronic participants’ ARAT scores with scores evaluating the degree of compensatory movements during ARAT, as derived from the study by Barth *et al*. [45], showing that compensatory behaviors were penalized in ARAT scoring. Similarly, the proposed biomarker exhibited significant negative correlations with compensatory movement scores, as shown in Extended Data Fig. 5B, suggesting that individuals exhibiting greater degrees of compensatory behavior in laboratory settings tended to achieve lower biomarker scores. We attribute this to the machine learning model’s design, which learned to identify meaningful phenotypes in motor performance that strongly correlated with clinician-evaluated scores across cross-sectional and longitudinal data.

## Discussion

In this study, we developed and validated a digital biomarker for post-stroke upper-limb motor recovery by aggregating kinematic characteristics of linear movements during natural behaviors using a single wrist-worn sensor. The proposed biomarker demonstrated excellent interpretability, reliability, concurrent validity, known-group validity, discriminant validity, and responsiveness, particularly in subacute patients. Additionally, through a Monte Carlo simulation framework, we showed that the biomarker could support more efficient clinical trials by leveraging its low measurement error compared to clinician-performed endpoints. The biomarker also displayed strong generalizability to chronic patients, further supporting its validity across different stages of chronicity.

This work builds on prior in-laboratory studies that investigated how kinematics of linear movements change with motor recovery, particularly in relation to the theory of submovements [17, 46]. These studies have shown that linear wrist movements performed by stroke survivors on a 2D tabletop become more fragmented with lower peak velocity and shorter displacement as severity increases [14–17]. With rehabilitation, their movements generally become faster and larger, showing higher peak velocities and greater displacement. These characteristics align with the features that constitute the proposed digital biomarker (e.g., features 2 and 4 in Table 2), indicating that displacement and velocity of linear movements increase with recovery.

Some prior studies have attempted to extend these in-laboratory findings to analyze linear movements during naturalistic upper-limb behaviors [18, 47]. However, these studies were conducted in simulated home environments rather than real-world settings and involved only brief periods of data collection (e.g., less than an hour per participant). Moreover, they primarily focused on demonstrating concurrent validity without assessing responsiveness or reliability. Despite these limitations, our findings resonate with these studies, underscoring the importance of variability in movement segment kinematics (features 1 and 3 in Table 2) as critical indicators of motor recovery [18, 19, 42].

The proposed biomarker is a measure of *motor performance* within the International Classification of Functioning, Disability, and Health (ICF) framework [48]. Despite this, it demonstrates strong cross-sectional and longitudinal associations with clinical measures that assess distinct constructs: FMA-UE for body function and structure, and ARAT for functional capacity [49]. This is because the proposed biomarker is designed to learn relevant features of motor performance, exhibiting strong cross-sectional and longitudinal associations with clinical measures. Unlike existing wearable-based measures, which are often limited to unidimensional characteristics of limb activity and may rely on two wrist sensors [13], the proposed biomarker provides a comprehensive view of relevant stroke recovery phenotypes only using a single sensor [41, 50].

This study has several limitations, highlighting important avenues for future research. Firstly, our data included only three-axis accelerometer data, requiring a high-pass filter to mitigate the effects of gravity. However, this approach assumes relatively constant wrist orientation during linear movements, an impractical assumption that may not accurately reflect patients’ actual movements. This limitation could introduce noise into the data features and, ultimately, the digital biomarker. Future studies should consider collecting both accelerometer and gyroscope data to accurately track linear acceleration in a global coordinate system [35]. Secondly, the dataset was limited to 24 hours of accelerometer data per clinical visit, so the ICC and SEM calculations provide estimates rather than actual reliability across day-to-day variations. Future work should collect data over extended periods, which may also help reduce measurement error and improve reliability, as suggested both theoretically [5] and empirically [10]. Lastly, the increased variability in the digital biomarker for a given ARAT score in chronic patients, as shown in Fig. 3, warrants further investigation. While we posit that individual-level factors may contribute to this variability [25], our study does not offer definitive evidence to support this. Future research could incorporate additional sensors or qualitative assessments of individual factors to better understand their impact on the biomarker in chronic patients.

In conclusion, we have developed and validated a digital biomarker to assess upper-limb motor recovery in stroke survivors using a single wrist-worn accelerometer, collecting data from patients’ naturalistic settings. The biomarker demonstrates strong validity, particularly among subacute-phase stroke survivors. These promising results support the biomarker’s potential as an endpoint in clinical trials, offering cost-effectiveness, low data collection burden, and reduced reliance on specialized clinicians its for large-scale deployment. It also holds promise for clinical practice, where it could help clinicians further personalize treatment strategies [2, 3] and enable patients to self-track and manage their rehabilitation progress [21].

## Methods

### Dataset

This study utilized measurement data from three separate research projects [25–29], publicly available through the NICHD DASH repository [23, 24]. The dataset included both subacute and chronic stroke patients as well as healthy subjects, as summarized in Fig. 1B.

For the subacute dataset, data were collected from 61 individuals across up to eight assessment sessions within the first six months post-stroke, totaling 224 sessions. These sessions were scheduled at weeks 2, 4, 6, 8, 12, 16, 20, and 24 from baseline. On average, subacute participants completed 3.67 ± 2.75 assessments. At the time of baseline assessment, they were 7.1 ± 3.1 days post-stroke. Participants received medical care as per their clinical plan. The chronic dataset consisted of measurement data from 79 stroke survivors at least six months post-stroke (on average 712 ± 1091 days). These patients participated in a clinical trial evaluating the effectiveness of an intervention, completing a total of 615 weekly assessments (7.77 ± 3.64 assessments per participant). Seventy-five percent of these patients received an eight-week intervention, while the remaining 25% extended their intervention by up to four weeks until reaching a plateau on the primary outcome measure (i.e., ARAT). The healthy dataset were obtained from 75 community-dwelling adults without any neurological conditions. They were assigned with the maximum clinical scores, without administrating the motor assessments, indicating maximal motor capacity and no impairment.

For all three groups, ActiGraph GTX3 accelerometer data were collected from both wrists for approximately 24 hours in their naturalistic environments following clinical assessment. For the subacute and chronic datasets, only accelerometer data from the stroke-affected limb were included in the analysis, whereas for the healthy dataset, data from both wrists were analyzed. The subacute dataset comprised 5,376 hours of accelerometer recordings, the chronic dataset included 13,860 hours, and the healthy dataset contained 3,600 hours of recordings. All raw accelerometry data were sampled at 30 Hz. A bandpass filter was applied between 0.25 and 2.5 Hz to approximate gravity-free acceleration and attenuate motion artifacts and inherent sensor noise for subsequent analysis [25, 26].

### Linear movement detection and segmentation

Our data analytic pipeline began by identifying linear movements from continuous gravity-free acceleration data. Linear movements are the most frequently performed and fundamental upper-limb actions required for ADLs [51]. In-laboratory studies have also demonstrated that linear movements contain rich kinematic characteristics that can effectively reflect motor recovery [16, 17]. As outlined in Algorithm 1, we employed a heuristic algorithm to detect linear movements with the following criteria: 1) signal variance is concentrated in a single eigenvector, indicating a relatively straight path in 3D space, determined by Principal Component Analysis (PCA) on the 3D gravity-adjusted accelerometer data, where the variance percentage of the first PC exceeds 82% [18, 52]; 2) the movement duration is between 1 and 5 seconds [53];and 3) the movement magnitude (i.e., activity index) must exceed 0.0045 *g* [53]. On average, we observed 306.9 ± 221.5 linear movements per hour in subacute stroke survivors, 315.2 ± 217.7 per hour in chronic stroke survivors, and 662.4 ± 142.3 per hour in healthy subjects.

After identifying linear movement episodes, we derived the velocity time-series by integrating acceleration data along the primary and secondary movement directions (i.e., the first two principal components derived earlier). Our analysis focused on the kinematic profiles within these dimensions, where the majority of signal variance was captured, thereby reducing both feature dimensionality and computational complexity. Movement segments were then identified based on zero-crossings in the velocity time-series for each direction [18, 47].

#### Algorithm 1

Linear movement identification

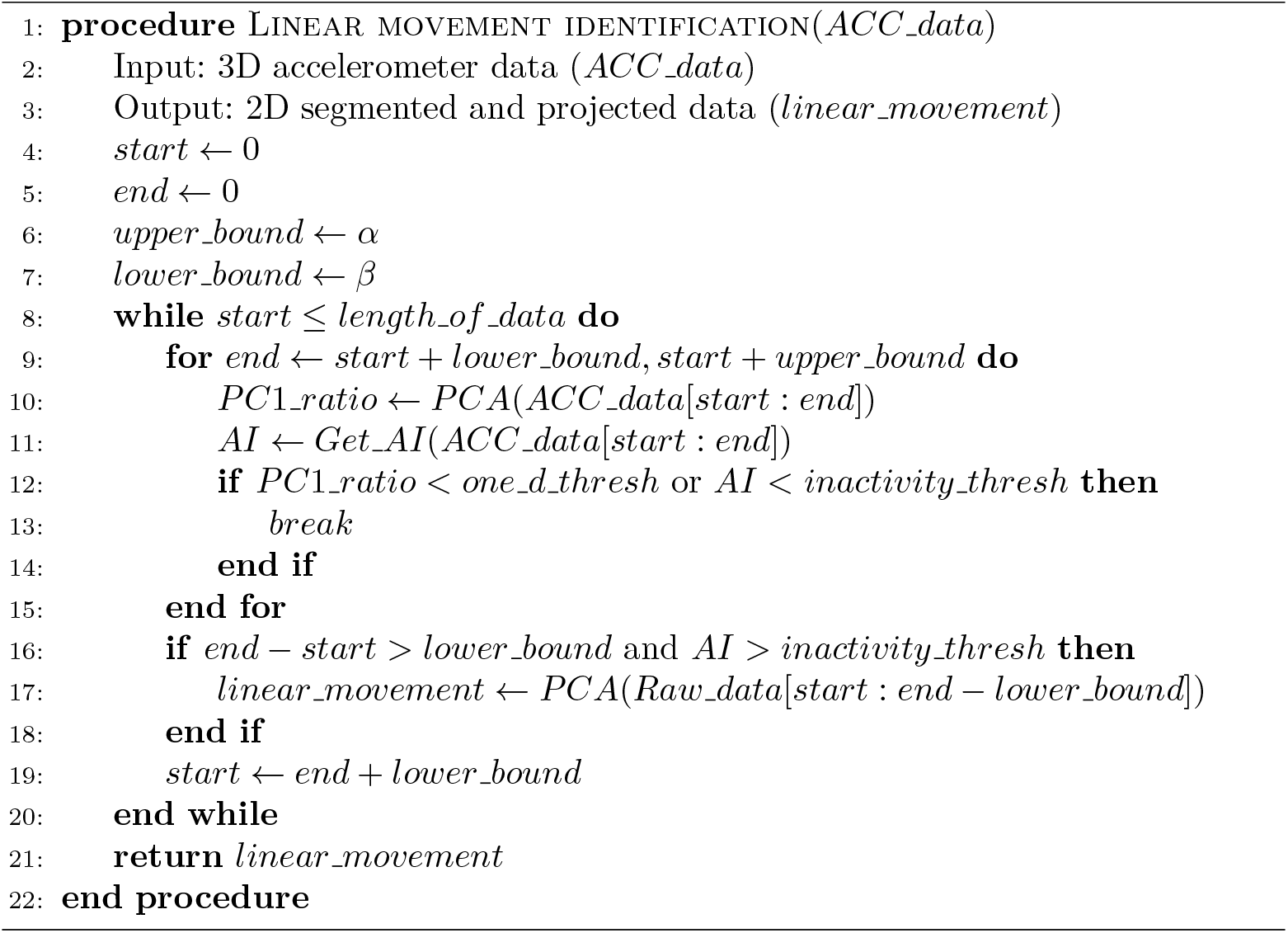

### Features extraction

Research indicates that movement segments contain unique kinematic and temporal patterns relevant to stroke recovery [18, 47], likely resulting from impairments in motor planning and control mechanisms, such as feedforward and feedback control [54, 55]. To capture these variations more precisely, movement segments were clustered into three subsets: the homogeneous set, the outlier set, and the entire set. The homogeneous and outlier sets were identified using the unsupervised clustering methods outlined in prior work [19]. This approach builds on findings that the velocity profiles of linear movement segments in neurologically intact individuals typically exhibit a smooth, bell-shaped curve [31, 56], whereas stroke survivors often display more irregular morphologies [16, 17]. The entire set encompassed all movement segments, combining the homogeneous and outlier sets. Data features were extracted independently from these subsets to capture nuanced, context-specific characteristics (i.e., homogeneous and outlier sets), as well as more comprehensive characteristics (i.e., entire set).

Data features encompassed kinematic, morphological, and temporal characteristics of movement segments in each cluster. Kinematic features were derived from descriptive statistics—such as minimum, maximum, mean, median, standard deviation, 10^th^ percentile, 90^th^ percentile—calculated on displacement, velocity, acceleration, and jerk profiles of each movement segment. For morphological features, we first resampled the velocity profiles of the segments to a normalized time duration and applied PCA to extract the top five basis functions, representing both high- and low-frequency movement components, as described in our prior study [30]. The aforementioned descriptive statistics were then applied to the values of these basis functions to capture the morphological characteristics of movement segments. Temporal features were designed to capture inter-segment dynamics [30], so we extracted them solely from the entire set of movement segments. A 2D histogram represented the probability density of consecutive segments, from which geometric features were derived to describe this distribution, including bounding box, convex hull, and eccentricity [57]. Descriptive statistics were then applied to the bounding box dimensions, and to the convex hull area, perimeter, compactness, and eccentricity. These kinematic, morphological, and temporal descriptive features were processed within each subject to summarize subject-level characteristics using aggregation functions, such as minimum, maximum, mean, median, standard deviation, 10^th^ percentile, 90^th^ percentile.

### Machine learning model

The training process was designed around the three key pillars of biomarker validation criteria: reliability, concurrent validity, and responsiveness. We first pruned unreliable and irrelevant features based on the following criteria: an ICC(3,1) *<* 0.75 [34], a Spearman’s correlation coefficient *<* 0.75 for cross-sectional correlations with ARAT or FMA-UE [36], a Spearman’s correlation coefficient *<* 0.4 between longitudinal changes in the feature and changes in clinical assessments [37], or an AUC-ROC *<* 0.7 for detecting responders based on the MDC of the clinical assessments [37]. To streamline training and prevent multicollinearity, inter-feature correlations were computed, and lower-performing features within highly correlated pairs (Pearson’s correlation *>* 0.9) were removed.

Next, multivariate linear mixed-effects models with L2 regularization were used to combine the remaining features into a composite biomarker. These models incorporated both fixed and random effects, where 1) fixed effects captured population-level correlations with the min-max-normalized ARAT score, addressing concurrent validity and 2) random effects accounted for individual-level variation, with an average of 3.67 ± 2.75 longitudinal data points per subject used for training, to ensure longitudinal consistency (i.e., responsiveness). Each subject was treated as a grouping factor to model individual-level data. The ARAT score was selected as the primary target variable because it was the only clinical measure available for chronic patients and strongly correlates with FMA-UE (Spearman’s ρ = 0.96 with *p* = 8.9 × 10^−120^ from our dataset) [58]. It is worth highlighting that we did not intend to use ARAT to devise a digital version of itself, as it is not a perfect gold standard for assessing motor recovery. Rather, we used it to capture associations, aligning with best practices in digital biomarker research [43]. With min-max normalization applied to the target clinical measure (ARAT), the primary range of the digital biomarker was from 0 to 1, although its theoretical range was unbounded.

The models were trained, validated, and tested using nested LOSOCV, ensuring no data from the same subject was used across these stages. The outer cross-validation was dedicated to testing, while the inner cross-validation optimized the L2 regularization parameter. This framework is essential for evaluating the generalizability of digital biomarkers [32].

## Evaluation

This study estimated the test-retest reliability of the digital biomarker to assess its consistency against potential day-to-day variability. Due to data constraints of only 24 hours per subject, we split the dataset into two groups by alternating the group categorizing every other 30-minute time window (i.e., alternating between even- and odd-numbered time windows). We assumed that a 30-minute window would be sufficient for individuals to engage in various activities, allowing us to assess the biomarker’s reliability across a diverse set of behaviors.

To quantify measurement error, we estimated the SEM and MDC. SEM represents the inherent error in a measure due to unreliability, while MDC estimates the smallest detectable score change that reflects a true difference, rather than random measurement error or variability [35]. SEM and MDC were calculated using

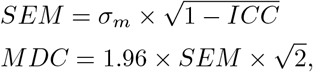

where *σ*_*m*_ is the standard deviation of outcome measures. To estimate SEM and MDC for ARAT and FMA-UE, *σ*_*m*_ was calculated from our dataset, while the ICC value reflecting inter-rater variability was referenced from existing literature (i.e., ICC of 0.965 for both ARAT and FMA-UE) [58]. This implies that, although the SEM and MDC comparisons were not entirely derived from the same dataset, they accounted for the primary sources of variability in the measurement: test-retest variability for the digital biomarker and inter-rater variability for clinical measures.

Concurrent validity examines the extent to which an outcome measure (i.e., digital biomarker) is an adequate reflection of a gold standard (e.g., FMA-UE and ARAT). In this study, we evaluated concurrent validity using Spearman’s correlation coefficients, given the non-linear relationships observed in Figs. 2 and 3. For known-group validity, the Wilcoxon rank-sum test was used to compare differences between stroke survivors and healthy subjects. Discriminant validity examines if the construct captured by the proposed biomarker was different from other constructs such as age, gender, and limb dominance in this study. The underlying hypotheses were the biomarker should not be influenced by motor characteristics associated with the aging process, gender, or limb dominance. We computed Spearman’s correlation between age and the proposed biomarker at baseline, and used the Wilcoxon rank sum test to compare the biomarker values at baseline between gender groups and between affected dominant and non-dominant limbs.

Responsiveness refers to the ability of a measure to detect meaningful changes over time in the construct being assessed. In our study, we validated the proposed biomarker using three approaches. First, we evaluated the correlation between the temporal trajectories of the proposed biomarker and standardized clinical assessments using Spearman’s correlation coefficient. Second, we computed the correlation of changes between the proposed biomarker and standard clinical assessments [37] to assess whether the biomarker could reflect changes in clinical assessments over the course of therapy. Lastly, we evaluated whether the proposed biomarker could detect improvement, as defined by standardized measures, using AUC-ROC [37]. Stroke survivors were categorized as “responders” or “non-responders” based on whether the change in standardized assessments between their first and last visits exceeded the computed MDC, as summarized in Table 1. AUC-ROC was then used to examine the agreement between the improvement in the proposed biomarker and the classification of patients as responders or non-responders.

### Hypothetical clinical trial

We implemented a hypothetical clinical trial using the statistical simulation framework proposed by Mori *et al*. [11]. The simulation was modeled after the randomized clinical trial conducted by Boake *et al*. [38], which compared the efficacy of constraint-induced movement therapy (CIMT) versus conventional therapy among subacute stroke survivors. The trial used the FMA-UE as the primary endpoint to evaluate the treatment effect between the two groups.

Population means and variances for changes in FMA-UE in both groups were derived from Boake *et al*.’s trial [38]. We estimated the population means for improvement in the proposed biomarker, assuming it would yield the same percentage of improvement as the FMA-UE, given the relatively linear relationship observed in Fig. 2B. However, since the wearable-based assessment exhibited significantly smaller SEM compared to the FMA-UE, the population variance was reduced proportionally for the proposed biomarker. The difference in reliability between the biomarker (ICC(3, 1) = 0.93) and the FMA-UE (ICC(3, 1) = 0.96 [58]) was negligible, and therefore its impact on the variance was disregarded for simplicity.

Monte Carlo simulation was performed by sampling means and variances from Gaussian and Chi-squared distributions, respectively. A two-tailed Welch’s t-test was used to account for unequal variances, and hypothesis support was defined as achieving statistical significance between the intervention and control groups with *α* = 0.05. The simulation iterated over 10,000 trials for sample sizes ranging from 2 to 250, calculating the probability of hypothesis support as the proportion of trials achieving significance. This framework estimated the minimum sample size required for each assessment to achieve adequate statistical power, highlighting the potential efficiency gains of the wearable-based digital biomarker.

## Data Availability

The data sued in this study could be found here https://dash.nichd.nih.gov/study/426315
https://dash.nichd.nih.gov/study/426433

## Extended Data

**Extended Data Fig 1.**
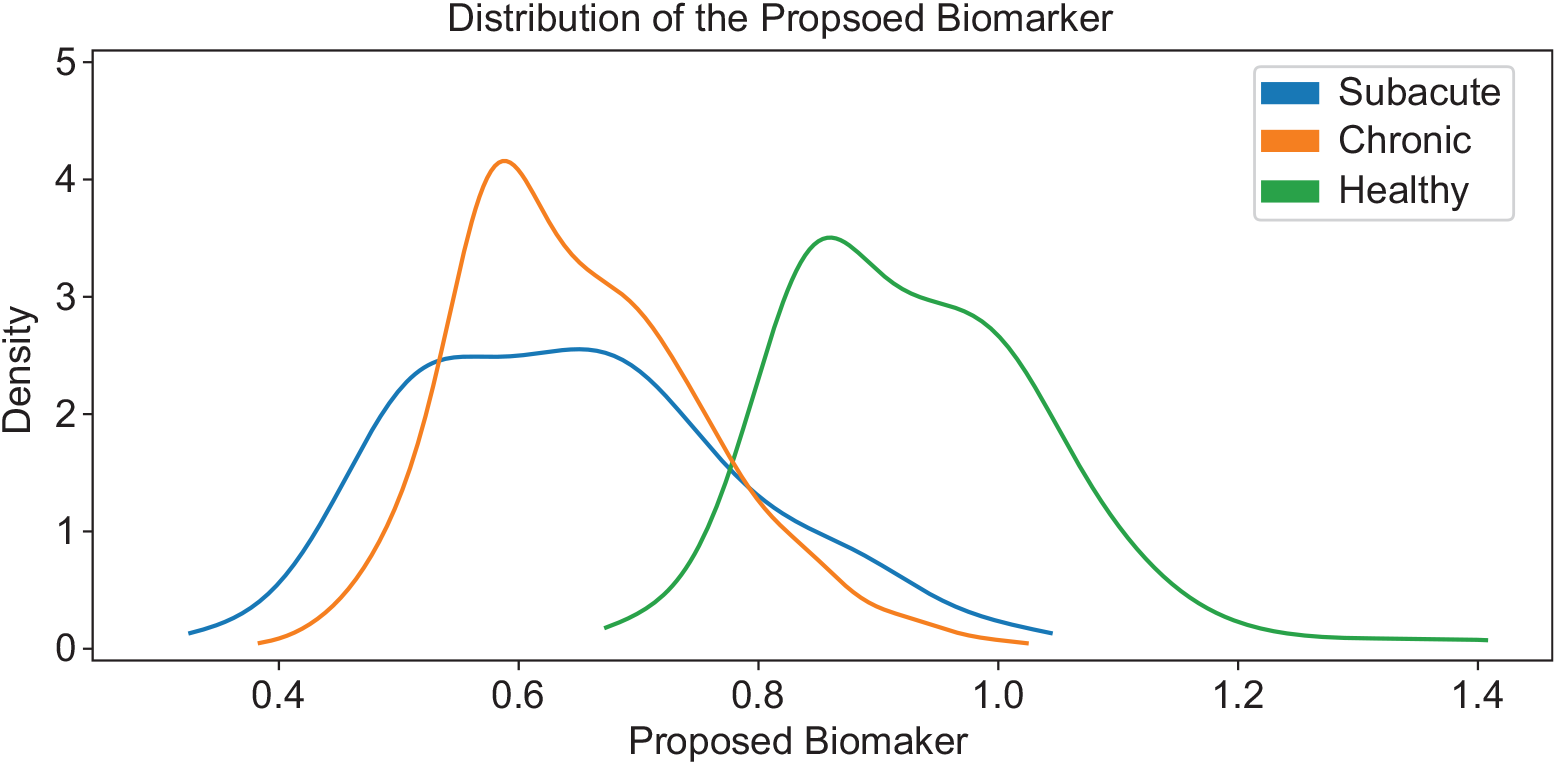
Distribution of the proposed biomarker. The plot showed the distribution of the proposed biomarker in healthy subjects (green), subacute (blue), and chronic (orange) stroke survivors. Kernel density estimation was used to visualize the distribution of the proposed biomarker.

**Extended Data Fig 2.**
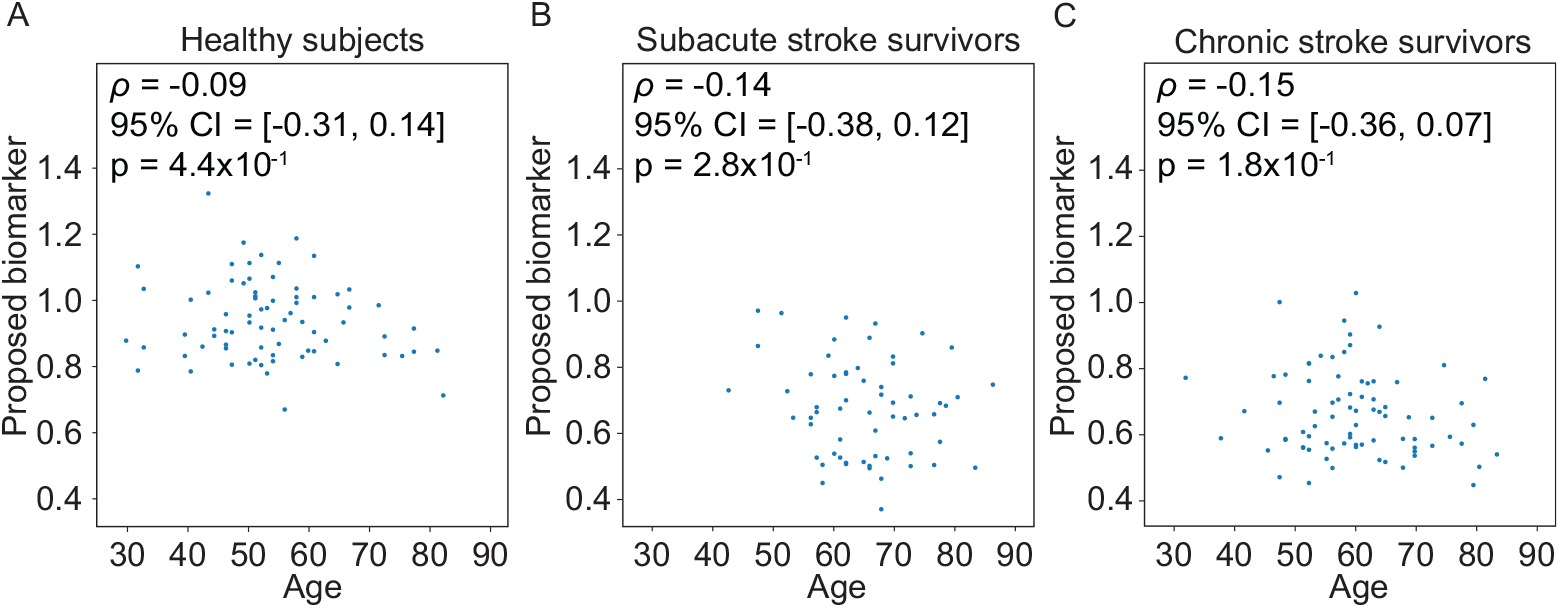
Discriminant validity in age. **A**-**C**. The scatter plots showed the relationship between age and the proposed biomarker at baseline for healthy, subacute, and chronic subjects respectively. Spearman’s correlation was used for computing correlation. For stroke survivors, only baseline data were included for comparison. For healthy subjects, data from only one wrist were used.

**Extended Data Fig 3.**
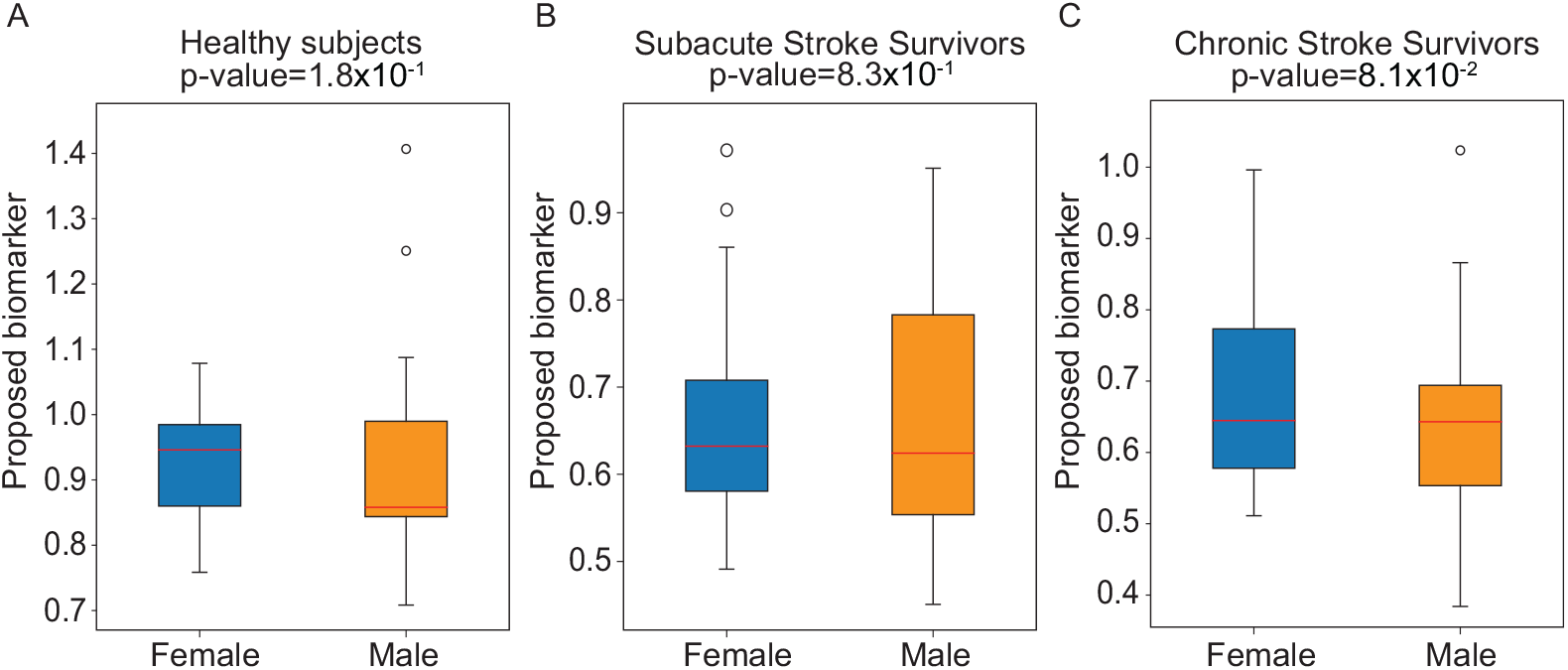
Discriminant validity in gender. **A**. The boxplot show the comparison of the proposed biomarker between female and male in healthy subjects. Wilcoxon rank sum test was used for evaluating significant difference. **B**. The boxplots show the comparison between female and male in proposed biomarker, ARAT, and FMA-UE in subacute survivors. **C**. The boxplots show the comparison between female and male in proposed biomarker and ARAT in chronic survivors. For stroke survivors, only baseline data were included for comparison. For healthy subjects, data from only one wrist were used.

**Extended Data Fig 4.**
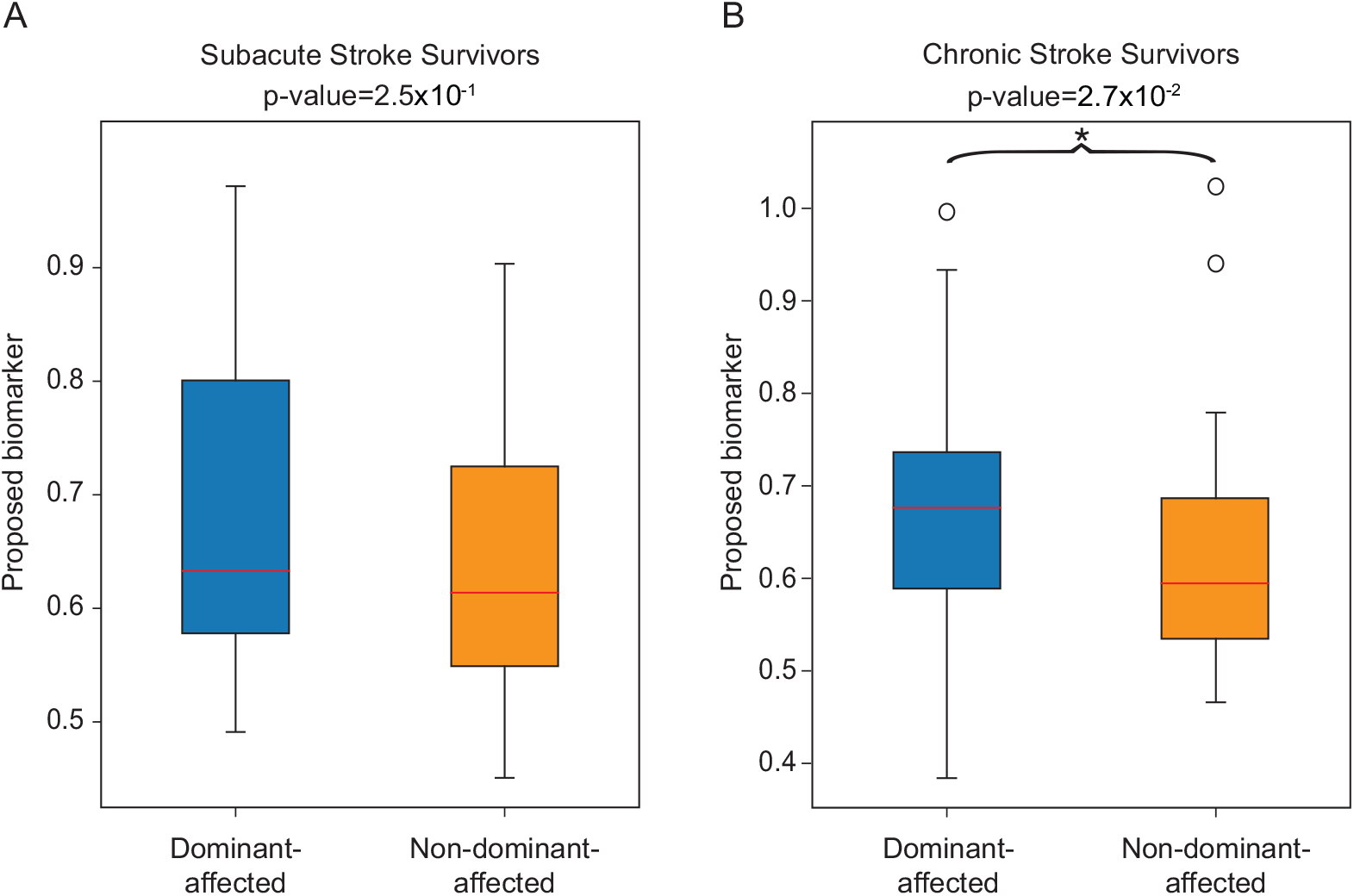
Discriminant validity in limb dominance. **A**. The boxplot shows a comparison of the proposed biomarker, ARAT, and FMA-UE between cases where the affected limb is the dominant limb and where it is not among subacute subjects. A Wilcoxon rank-sum test was used to evaluate significant differences. **B**. The boxplot shows a comparison of the proposed biomarker and ARAT between cases where the affected limb is the dominant limb and where it is not among chronic subjects. For stroke survivors, only baseline data were included for comparison.

**Extended Data Fig 5.**
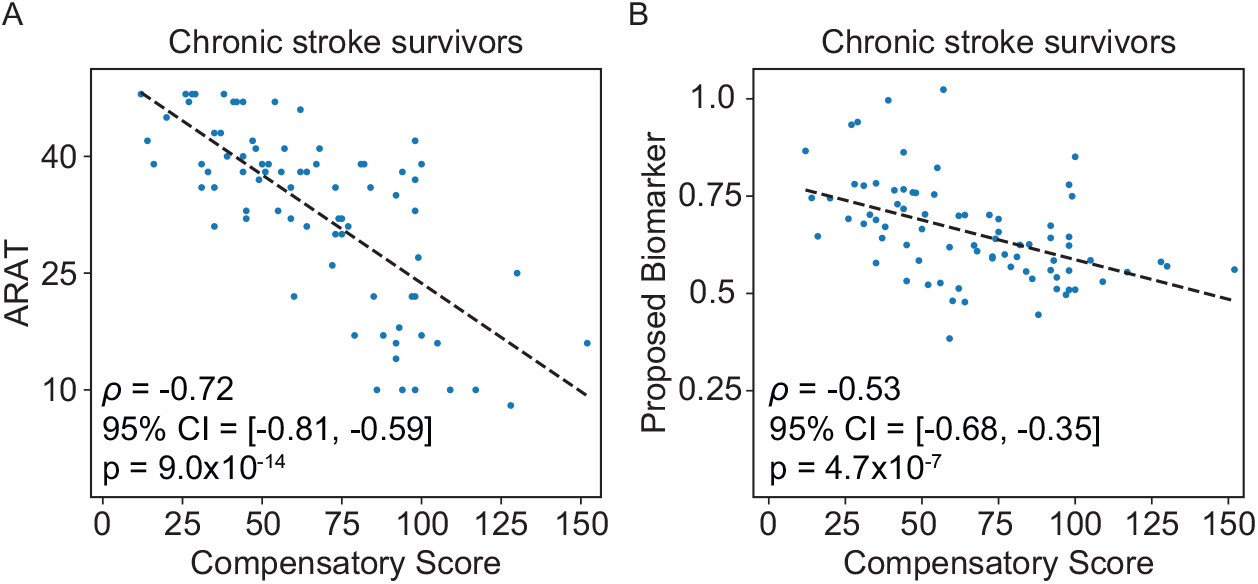
Relationship between compensatory scores, ARAT, and the proposed biomarker of chronic survivors. **A** The scatter plot illustrates the relationship between compensatory scores and ARAT assessment. Spearman’s correlation was used to assess the association between compensatory scores and ARAT assessment. **B** The scatter plot illustrates the relationship between compensatory scores and the proposed biomarker. Spearman’s correlation was used to assess the association between compensatory scores and the proposed biomarker.

## References

[1] Ali, M., English, C., Bernhardt, J., Sunnerhagen, K. & Brady, M. More outcomes than trials: a call for consistent data collection across stroke rehabilitation trials. International Journal of stroke 8, 18–24 (2013).

[2] Cramer, S. C. et al. The utility of domain-specific end points in acute stroke trials. Stroke 52, 1154–1161 (2021).

[3] Adans-Dester, C. P., Lang, C. E., Reinkensmeyer, D. J. & Bonato, P. Wearable sensors for stroke rehabilitation. Neurorehabilitation Technology 467–507 (2022).

[4] Rabadi, M. H. & Rabadi, F. M. Comparison of the action research arm test and the fugl-meyer assessment as measures of upper-extremity motor weakness after stroke. Archives of physical medicine and rehabilitation 87, 962–966 (2006).

[5] De Vet, H. C., Terwee, C. B., Mokkink, L. B. & Knol, D. L. Measurement in Medicine: A practical guide (Cambridge University Press, 2011).

[6] Adans-Dester, C. et al. Enabling precision rehabilitation interventions using wearable sensors and machine learning to track motor recovery. NPJ digital medicine 3, 121 (2020).

[7] See, J. et al. A standardized approach to the fugl-meyer assessment and its implications for clinical trials. Neurorehabilitation and neural repair 27, 732–741 (2013).

[8] Desowska, A. & Turner, D. L. Dynamics of brain connectivity after stroke. Reviews in the Neurosciences 30, 605–623 (2019).

[9] Ricotti, V. et al. Wearable full-body motion tracking of activities of daily living predicts disease trajectory in duchenne muscular dystrophy. Nature medicine 29, 95–103 (2023).

[10] Haberkamp, M. et al. European regulators’ views on a wearable-derived performance measurement of ambulation for duchenne muscular dystrophy regulatory trials. Neuromuscular Disorders 29, 514–516 (2019).

[11] Mori, H., Wiklund, S. J. & Zhang, J. Y. Quantifying the benefits of digital biomarkers and technology-based study endpoints in clinical trials: project moneyball. Digital Biomarkers 6, 36–46 (2022).

[12] Uswatte, G. et al. Objective measurement of functional upper-extremity movement using accelerometer recordings transformed with a threshold filter. Stroke 31, 662–667 (2000).

[13] Hayward, K. S. et al. Exploring the role of accelerometers in the measurement of real world upper-limb use after stroke. Brain Impairment 17, 16–33 (2016).

[14] Miranda, J. G. V. et al. Complex upper-limb movements are generated by combining motor primitives that scale with the movement size. Scientific reports 8, 12918 (2018).

[15] Krebs, H. I., Aisen, M. L., Volpe, B. T. & Hogan, N. Quantization of continuous arm movements in humans with brain injury. Proceedings of the National Academy of Sciences 96, 4645–4649 (1999).

[16] Rohrer, B. et al. Movement smoothness changes during stroke recovery. Journal of neuroscience 22, 8297–8304 (2002).

[17] Rohrer, B. et al. Submovements grow larger, fewer, and more blended during stroke recovery. Motor control 8, 472–483 (2004).

[18] Oubre, B. & Lee, S. I. Detection and assessment of point-to-point movements during functional activities using deep learning and kinematic analyses of the stroke-affected wrist. IEEE Journal of Biomedical and Health Informatics 28, 1022–1030 (2024).

[19] Oubre, B. et al. Estimating upper-limb impairment level in stroke survivors using wearable inertial sensors and a minimally-burdensome motor task. IEEE Transactions on Neural Systems and Rehabilitation Engineering 28, 601–611 (2020).

[20] FDA-NIH Biomarker Working Group. BEST (Biomarkers, Endpoints, and Other Tools) resource - Monitoring Biomarker. https://www.ncbi.nlm.nih.gov/books/NBK402282/. Accessed: 2024-05-13.

[21] Sahely, A., Giles, D., Sintler, C., Soundy, A. & Rosewilliam, S. Self-management interventions to improve mobility after stroke: an integrative review. Disability and rehabilitation 45, 9–26 (2023).

[22] Miller, A. E. et al. A large harmonized upper and lower limb accelerometry dataset: A resource for rehabilitation scientists. medRxiv (2024).

[23] Lang, C. Harmonized upper and lower limb accelerometry data part1 (version 1). NICHD Data and Specimen Hub (2024). URL 10.57982/fayx-p832.

[24] Lang, C. Harmonized upper and lower limb accelerometry data part2 (version 1). NICHD Data and Specimen Hub (2024). URL 10.57982/72z7-m179.

[25] Lang, C. E. et al. Upper limb performance in daily life approaches plateau around three to six weeks post-stroke. Neurorehabilitation and neural repair 35, 903–914 (2021).

[26] Lang, C. E. et al. Dose response of task-specific upper limb training in people at least 6 months poststroke: a phase ii, single-blind, randomized, controlled trial. Annals of neurology 80, 342–354 (2016).

[27] Waddell, K. J. et al. Does task-specific training improve upper limb performance in daily life poststroke? Neurorehabilitation and neural repair 31, 290–300 (2017).

[28] Bailey, R. R., Klaesner, J. W. & Lang, C. E. An accelerometry-based methodology for assessment of real-world bilateral upper extremity activity. PloS one 9, e103135 (2014).

[29] Bailey, R. R. & Lang, C. E. Upper extremity activity in adults: referent values using accelerometry. Journal of rehabilitation research and development 50, 1213 (2014).

[30] Oubre, B. et al. Decomposition of reaching movements enables detection and measurement of ataxia. The Cerebellum 1–12 (2021).

[31] Flash, T. & Hogan, N. The coordination of arm movements: an experimentally confirmed mathematical model. Journal of neuroscience 5, 1688–1703 (1985).

[32] Hendrix, S. B. et al. Perspectives on statistical strategies for the regulatory biomarker qualification process. Biomarkers in Medicine 15, 669–684 (2021).

[33] Baker, K., Cano, S. J. & Playford, E. D. Outcome measurement in stroke: a scale selection strategy. Stroke 42, 1787–1794 (2011).

[34] Koo, T. & Li, M. A guideline of selecting and reporting intraclass correlation coefficients for reliability research. j chiropr med. 2016; 15 (2): 155–63 (2000).

[35] Weir, J. P. Quantifying test-retest reliability using the intraclass correlation coefficient and the sem. The Journal of Strength & Conditioning Research 19, 231–240 (2005).

[36] Bardy, T. L. The swiss health insurance literacy measure (hilm-ch): measurement properties and cross-cultural validation. BMC Health Services Research 23, 85 (2023).

[37] Sharma, S. et al. Reliability, validity, responsiveness, and minimum important change of the stair climb test in adults with hip and knee osteoarthritis. Arthritis care & research 75, 1147–1157 (2023).

[38] Boake, C. et al. Constraint-induced movement therapy during early stroke rehabilitation. Neurorehabilitation and neural repair 21, 14–24 (2007).

[39] Lee, S. I. et al. A novel upper-limb function measure derived from finger-worn sensor data collected in a free-living setting. PloS one 14, e0212484 (2019).

[40] Grattan, E. S., Velozo, C. A., Skidmore, E. R., Page, S. J. & Woodbury, M. L. Interpreting action research arm test assessment scores to plan treatment. OTJR: occupation, participation and health 39, 64–73 (2019).

[41] Han, C. E., Arbib, M. A. & Schweighofer, N. Stroke rehabilitation reaches a threshold. PLoS computational biology 4, e1000133 (2008).

[42] Okita, S., De Lucena, D. S. & Reinkensmeyer, D. J. Movement diversity and complexity increase as arm impairment decreases after stroke: Quality of movement experience as a possible target for wearable feedback. IEEE Transactions on Neural Systems and Rehabilitation Engineering (2024).

[43] Committee for Medicinal Products for Human Use. Draft qualification opinion for stride velocity 95th centile as primary endpoint in studies in ambulatory duchenne muscular dystrophy studies. Eur Med Agency 2–166 (2023).

[44] Harris, J. E. & Eng, J. J. Individuals with the dominant hand affected following stroke demonstrate less impairment than those with the nondominant hand affected. Neurorehabilitation and neural repair 20, 380–389 (2006).

[45] Barth, J., Klaesner, J. W. & Lang, C. E. Relationships between accelerometry and general compensatory movements of the upper limb after stroke. Journal of neuroengineering and rehabilitation 17, 1–10 (2020).

[46] Milner, T. E. A model for the generation of movements requiring endpoint precision. Neuroscience 49, 487–496 (1992).

[47] Lee, S. I. et al. Wearable-based kinematic analysis of upper-limb movements during daily activities could provide insights into stroke survivors’ motor ability. Neurorehabilitation and neural repair 38, 659–669 (2024).

[48] World Health Organization. International classification of functioning, disability and health: Icf. http://www.who.int/classifications/icf/en/)(2001.

[49] Lohse, K. R., Miller, A. E., Bland, M. D.Lee, J.-M. & Lang, C. E. Validation of real-world actigraphy to capture post-stroke motor recovery. medRxiv 2024–11 (2024).

[50] Dorsey, E. R., Venuto, C., Venkataraman, V., Harris, D. A. & Kieburtz, K. Novel methods and technologies for 21st-century clinical trials: a review. JAMA neurology 72, 582–588 (2015).

[51] Schambra, H. M. et al. A taxonomy of functional upper extremity motion. Frontiers in neurology 10, 857 (2019).

[52] Lee, J., Oubre, B., Daneault, J.-F., Lee, S. I. & Gupta, A. S. Estimation of ataxia severity in children with ataxia-telangiectasia using ankle-worn sensors. Journal of Neurology 270, 5097–5101 (2023).

[53] Bai, J. et al. An activity index for raw accelerometry data and its comparison with other activity metrics. PloS one 11, e0160644 (2016).

[54] Wu, C.-y., Trombly, C. A., Lin, K.-c. & Tickle-Degnen, L. A kinematic study of contextual effects on reaching performance in persons with and without stroke: influences of object availability. Archives of physical medicine and rehabilitation 81, 95–101 (2000).

[55] Rizzo, J.-R. et al. Motor planning poststroke: impairment in vector-coded reach plans. Physiological reports 3, e12650 (2015).

[56] Hoff, B. A model of duration in normal and perturbed reaching movement. Biological Cybernetics 71, 481–488 (1994).

[57] Mingqiang, Y., Kidiyo, K., Joseph, R. et al. A survey of shape feature extraction techniques. Pattern recognition 15, 43–90 (2008).

[58] Platz, T. et al. Reliability and validity of arm function assessment with standardized guidelines for the fugl-meyer test, action research arm test and box and block test: a multicentre study. Clinical rehabilitation 19, 404–411 (2005).

